# Lactate Dehydrogenase-to-Lymphocyte Ratio Predicts Early Mortality in Patients with Severe Fever with Thrombocytopenia Syndrome

**DOI:** 10.1101/2025.07.10.25331251

**Authors:** Ruize Ma, Jingxia Wang, Ranran Wang, Ruihua Zhang, Xiaoyu Xue, Yameng Mu, Hong xiao Wu, Ling Lin, Zhihai Chen

**Affiliations:** National Key Laboratory of Intelligent Tracking and Forecasting for Infectious Diseases, Beijing Ditan Hospital, Capital Medical University, Beijing, China; Department of Infectious Disease, Peking University Ditan Teaching Hospital, Beijing, China; Department of Infectious Diseases, Yantai City Hospital for Infectious Disease, Yantai, China

**Keywords:** Severe fever with thrombocytopenia syndrome, predictor, lactate dehydrogenase-to-lymphocyte ratio, predict model, nomogram

## Abstract

**Background:** Severe fever with thrombocytopenia syndrome (SFTS) is caused by SFTS virus (SFTSV) with high mortality.

**Methods:** Between January 2022 and October 2024, a total of 208 cases from Yantai Qishan Hospital have been included in this study. The sLLR was defined as the standardized ratio of lactate dehydrogenase to absolute lymphocyte count. Firth penalized regression was utilized for variable selection. The receiver operating characteristic (ROC) analysis compared sLLR with C-reactive protein/albumin ratio (CAR). A nomogram based on sLLR was validated through 1,000 bootstraps. Performance was assessed using AUC, Brier score, and decision curve analysis (DCA).

**Results:** Non-survivors had a higher sLLR level (median 2.88 vs. 1.15; P<0.001). sLLR showed superior discrimination compared to CAR (AUC 0.804 vs. 0.648, P=0.016), with 95.7% sensitivity. The nomogram achieved AUC 0.87 (95%CI 0.798-0.942) and validation AUC 0.876 (Brier score 0.074; Hosmer-Lemeshow P=0.778).

**Conclusions:** sLLR can be utilized as an independent prognostic factor for SFTS patients. The nomogram enables rapid mortality risk stratification, providing a practical tool for frontline clinicians in resource-limited settings.

**Synopsis:** This study identifies the standardized lactate dehydrogenase-to-lymphocyte ratio (sLLR) as a novel prognostic biomarker for early mortality in patients with Severe Fever with Thrombocytopenia Syndrome (SFTS). Through a retrospective cohort of 208 laboratory-confirmed SFTS cases, non-survivors exhibited significantly higher sLLR levels (median 2.88 vs. 1.15, P<0.001) compared to survivors. The sLLR demonstrated superior discriminatory power (AUC 0.804) over the established C-reactive protein/albumin ratio (CAR; AUC 0.648, P=0.016), with 95.7% sensitivity at a cutoff of 1.457. Multivariate Firth penalized regression confirmed sLLR as an independent predictor of mortality (OR=1.84, 95% CI 1.15–3.06; P=0.012), alongside neurological manifestations (OR=4.25). A clinically practical nomogram integrating sLLR, activated partial thromboplastin time (APTT), and neurological symptoms achieved robust performance (AUC 0.876 after 1,000 bootstraps) and calibration (Brier score 0.074), enabling rapid risk stratification in resource-limited settings. These findings illuminate sLLR’s pathophysiological relevance—reflecting virally driven tissue damage (elevated LDH) and immune depletion (lymphocytopenia)—and provide a tool to guide early intervention for high-risk SFTS patients.

## Introduction

Severe fever with thrombocytopenia syndrome (SFTS) caused by the SFTSV (Dabie bandavirus, family Phenuiviridae), is an acute tick-borne disease with a mortality rate of 12%-30%[1, 2]. It was first reported in China in 2009 and has since become endemic to East Asia[3–6]. Recently, homologous viruses have been detected in the United States[7], India[8] and Australia[9], indicating potential global spread[10]. The early clinical presentation of SFTS often lacks distinguishing characteristics, with initial symptoms that are similar but may progress into significantly varied clinical outcomes. Epidemiological data indicates that cases are predominantly distributed in rural mountainous regions within endemic areas, where healthcare infrastructure is often less advanced compared to urban centers[11]. The backward medical conditions and rapid clinical progression increase the burden on primary diagnosis and treatment management. There is no effective and specific therapy for SFTS. Therefore, it is crucial for rural primary care physicians to identify patients with life-threatening conditions as early as possible. Although supportive treatment has made progress, early identification of high-risk patients remains challenging.

SFTSV infection could involve direct viral cytotoxicity and dysregulation of host immune responses. Recent studies show that SFTSV damages the hematopoietic microenvironment by infecting endothelial cells, causing widespread cell rupture and releasing lactate dehydrogenase (LDH), which correlated with tissue damage[12]. Additionally, the virus induces apoptosis of CD4+ T lymphocytes and bone marrow suppression, leading to a decrease in lymphocyte count (LY)[13–15]. Theoretically, the ratio of LDH to LY can more sensitively reflect the severity of SFTS due to its reverse fluctuation trend in the early stage of the disease. Therefore, we propose the ratio of LDH to LY (LDH/LY) as a biomarker that can reflect the degree of cellular damage and immune suppression, illustrating the pathophysiological features of multiorgan damage in SFTS. Additionally, as a novel inflammatory biomarker, the ratio of LDH to LY has been validated for its significant role in the diagnosis and prognosis prediction of COVID-19[16]. Beyond the field of infectious diseases, the ratio of lactate dehydrogenase to lymphocytes has also been used as an important marker for assessing the prognosis of lymphoma patients[17]. However, the independent prognostic value of this ratio in SFTS and its role in risk stratification models remain unclear. In this study, we established a standardized ratio of LDH/LY (divided by 1000) (sLLR), aiming to evaluate the predictive value of sLLR for SFTS mortality and to develop a simple and feasible nomogram for early severity stratification and prediction of death outcomes.

## Methods

### Ethics Statement

This study was approved by the Ethics Committee of Beijing Ditan Hospital, Capital Medical University (Approval No.DTEC-KY2022-022-01) and was conducted in compliance with the principles outlined in the Declaration of Helsinki. Written informed consent from each patient was waived due to the study’s retrospective nature.

### Study Design and Population

This single-center retrospective cohort study enrolled 251 patients diagnosed with SFTS at the Yantai Qishan Hospital between January 2022 and October 2024. All patients should follow at least one of the following diagnostic criteria: (1) positive serum SFTSV RNA detection, (2) a ≥4-fold increase in antibody titers or seroconversion in paired serum samples collected ≥2 weeks apart, or (3) successful SFTSV isolation via cell culture. Based on predefined exclusion criteria, 43 patients were excluded due to: (1) coinfection with other viruses, (2) absence of laboratory-confirmed SFTS diagnosis, (3) incomplete clinical data, or (4) voluntary discharge or transfer to other hospitals.

### Data Collection

We systematically collected information related to patient demographic characteristics (age, sex, hospitalization, ICU admission, and clinical outcomes), comorbidities, symptoms, physical examination findings (including neurological manifestations), and laboratory tests (including blood routine, biochemical markers, kidney injury markers, coagulation, tissue damage markers, infection-related markers, and composite indicators). The observation endpoint of this study was defined as death or survival.

### Definitions

Neurological manifestations require documentation of at least one symptom, including headache, dizziness, consciousness disorder, apathy, bradyphrenia, somnolence, agitation, tremor, shivering, convulsions, or sluggish pupillary light reflex. Mucosal hemorrhagic manifestations included petechiae, purpura, ecchymosis, and rashes.

### Statistical Analysis

The sLLR was calculated as the ratio of lactate dehydrogenase (LDH, U/L) to lymphocyte count (LY, ×10⁹/L), standardized by dividing by 1,000. The categorical variables were expressed as frequency (%) using the χ2 test or Fisher’s exact test; normally distributed continuous variables were expressed as mean ± standard deviation (X±SD) using the T-test; and non-normal variables were expressed as median (interquartile range) using the Mann-Whitney U or Kruskal-Wallis test. Potential predictors of mortality were screened using univariate logistic regression analysis (P<0.2). To address the limited number of mortality events (n=23), Firth penalized regression was used to reduce bias in odds ratio estimation and construct a multivariate logistic model to calculate the risk ratio (OR) and 95% confidence interval. Multicollinearity was rigorously evaluated using Pearson’s correlation matrix (absolute coefficient threshold <0.8) and variance inflation factors (VIF <2). The goodness of fit of the model was tested using the likelihood ratio. The Box-Tidwell test was used to verify the linearity of the model. Model performance was evaluated through discrimination (AUC from ROC curves), calibration (Brier score, Hosmer-Lemeshow test, and GAM-smoothed calibration curve), and clinical utility (DCA net benefit). Internal validation was performed via 1,000 bootstrap resamples with replacement to assess model stability. Statistical analyses were performed using R 4.4.1 (Logistic, Firth, column-line graphs, DCA) and SPSS 27.0.1, with significance defined as a two-tailed P<0.05.

## Results

### Baseline Characteristics

Following the inclusion and exclusion criteria, a total of 208 patients with laboratory-confirmed SFTS were analyzed. Of these patients, 185 (88.9%) survived and 23 (11.1%) did not survive. Demographic information, clinical manifestation, and laboratory results are summarized in Table 1 and 2.

**Table 1.**
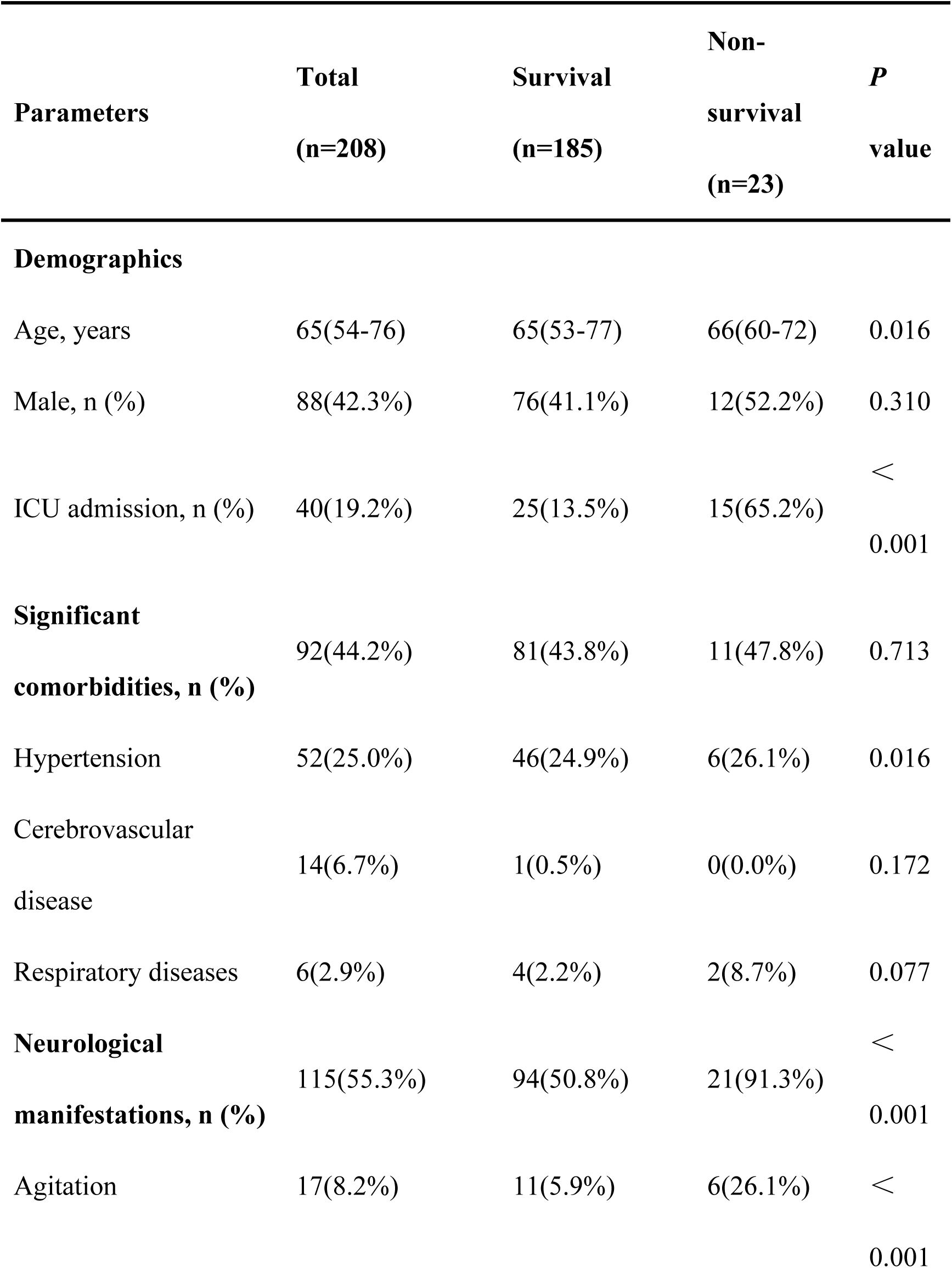
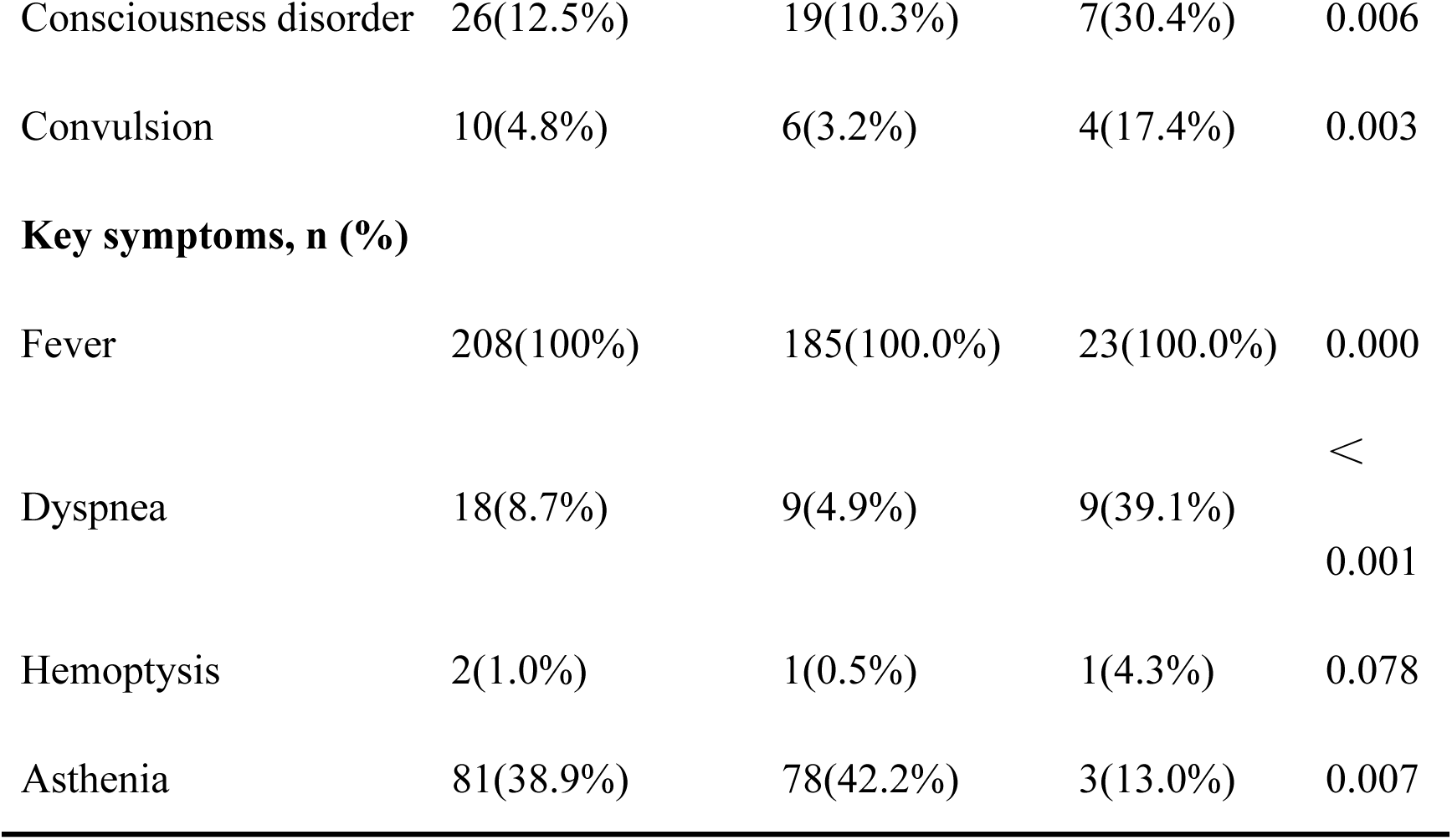
Baseline characteristics of SFTS patients stratified by survival status.

**Table 2.**
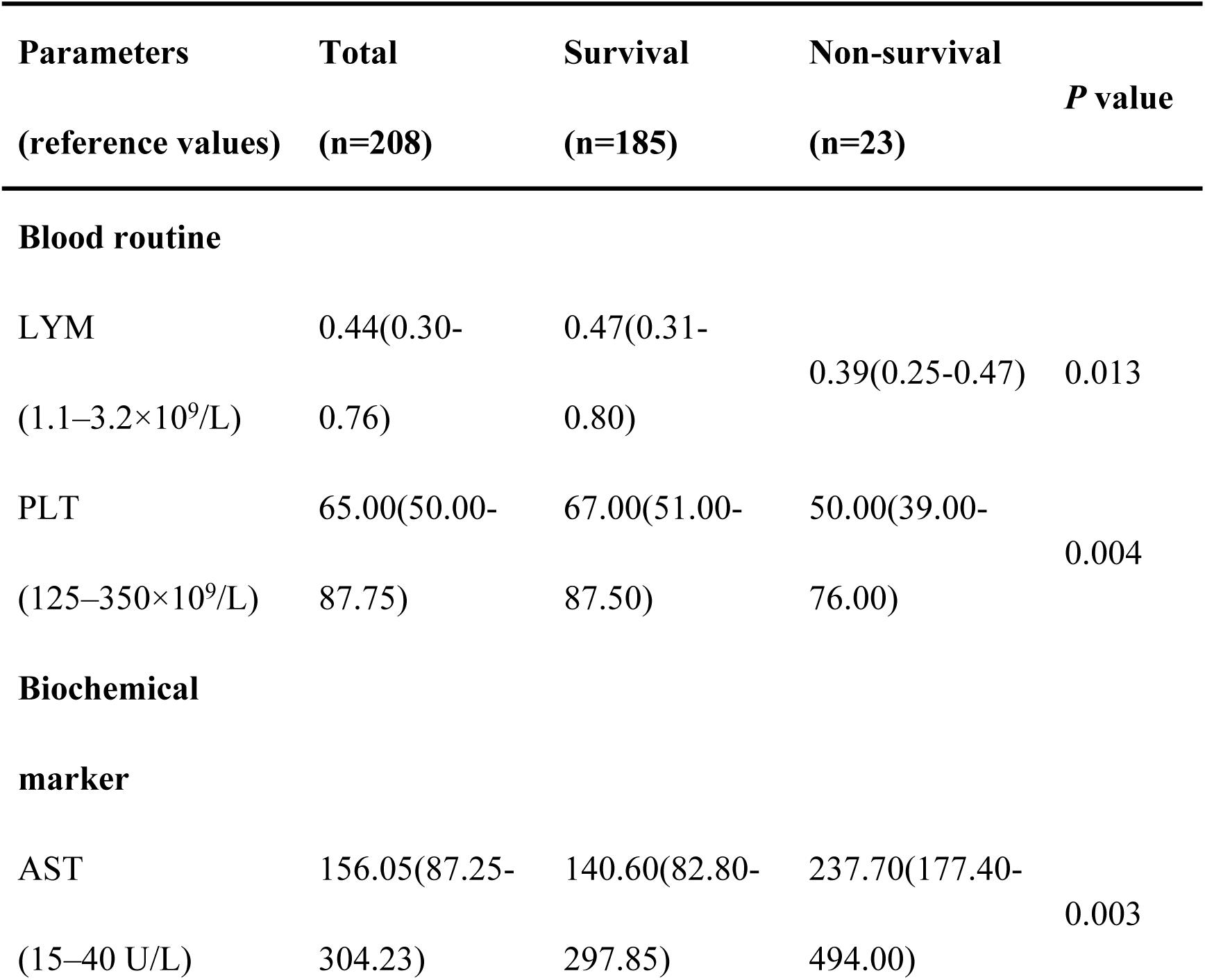
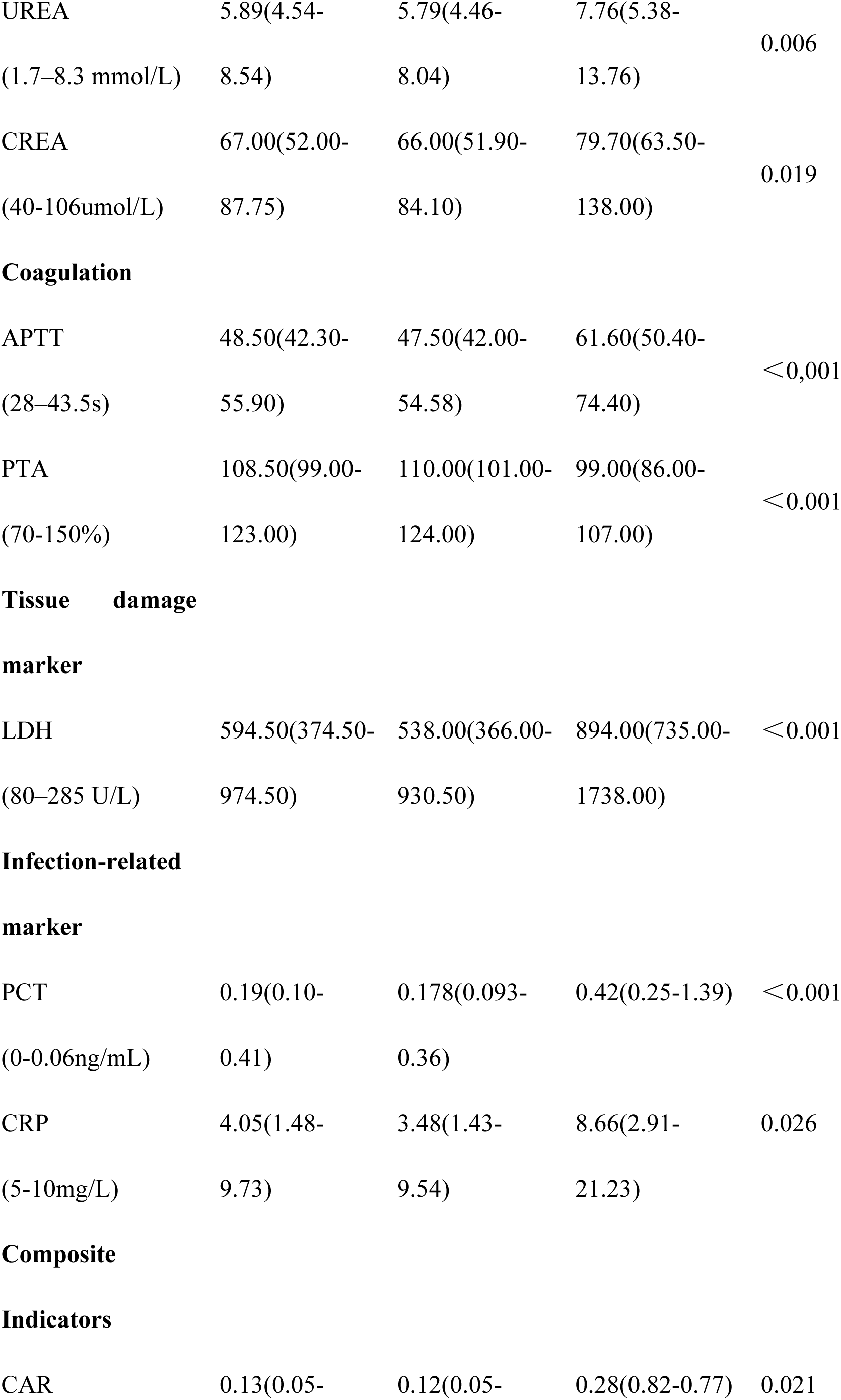
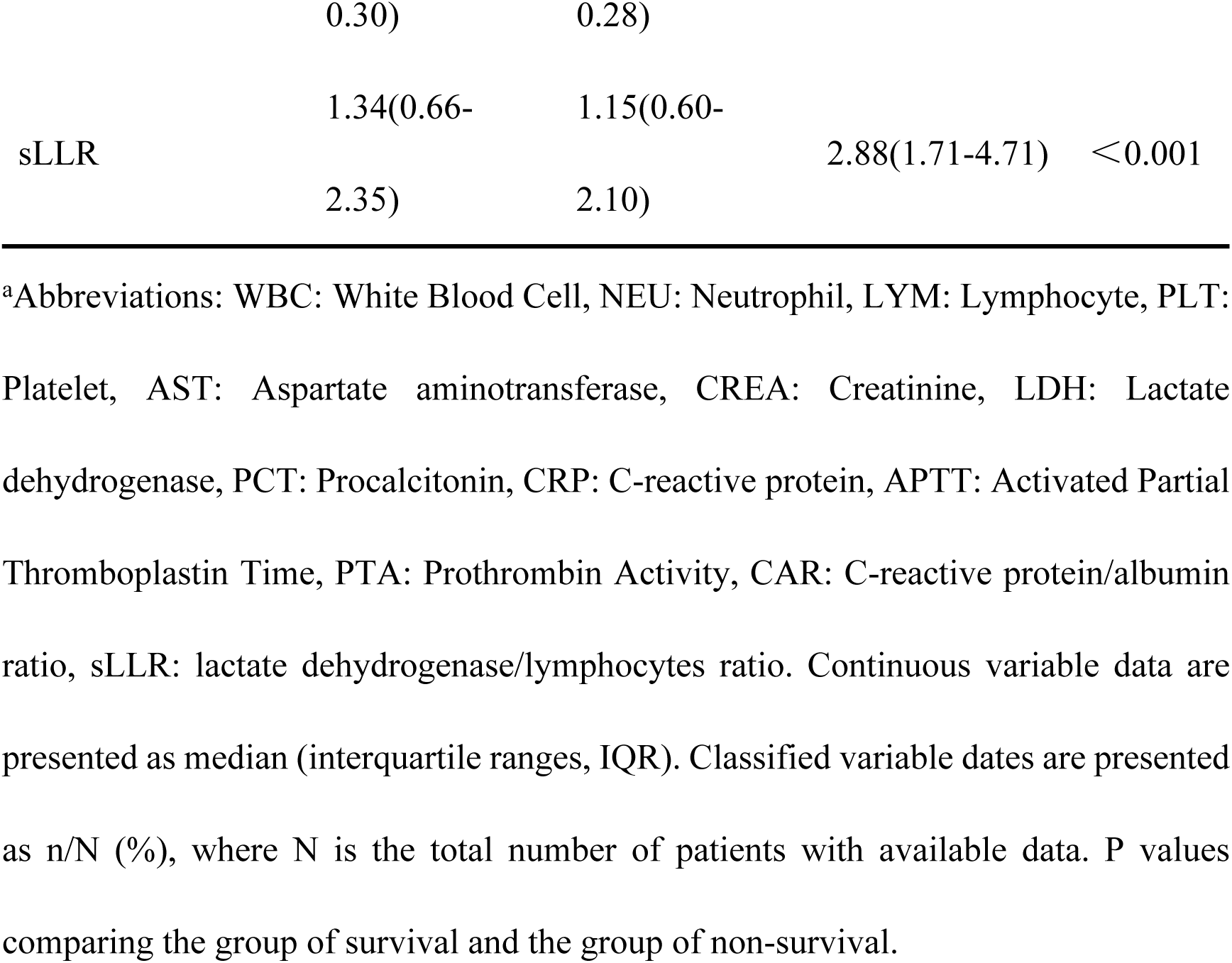
Laboratory results of patients with SFTS on admission.

The enrolled patients were divided into a survival group and a deceased group according to clinical outcome. Non-survivors were older than survivors and had a higher ICU admission rate and more frequent neurological manifestations. Specific neurological symptoms showed differences, including disturbance of consciousness, agitation, and convulsion.

Hematological analysis revealed lower lymphocyte counts in non-survivors compared to survivors, as well as lower platelet counts. Biochemical markers, including aspartate aminotransferase (AST), urea, and creatinine (CREA), were elevated in non-survivors.

Coagulation tests showed a prolonged activated partial thromboplastin time (APTT) in non-survivors. Additionally, tissue damage markers, notably lactate dehydrogenase (LDH), as well as infection-related indices including procalcitonin (PCT) and C-reactive protein (CRP), were markedly elevated in the deceased group. The sLLR was significantly higher in non-survivors (2.88 vs. 1.15, P < 0.001), could prove to be a more predictive marker than the CAR (0.28 vs. 0.12, P = 0.021).

### Univariate and Multivariate Analysis

Univariate logistic regression analysis showed that 17 variables might be associated with the risk of death in SFTS patients (P < 0.2), including neurological manifestations, sLLR, APTT and LDH, etc (Table 3). Firth penalized regression analysis retained two independent predictors: neurological manifestations (OR=4.24, 95% CI 1.01–27.05; P=0.048) and sLLR (OR=1.84, 95% CI 1.15–3.06; P=0.012). APTT prolongation showed a trend related to an increased risk of mortality (OR = 1.05, 95% CI 1.00–1.11, P = 0.071). The extremely high OR for the CAR indicated potential issues with multicollinearity or data sparsity. We observed that each 1-unit increase in sLLR corresponded to an 84% higher mortality risk, whereas patients with neurological manifestations had a 3.24-fold increased mortality risk.

**Table 3.**
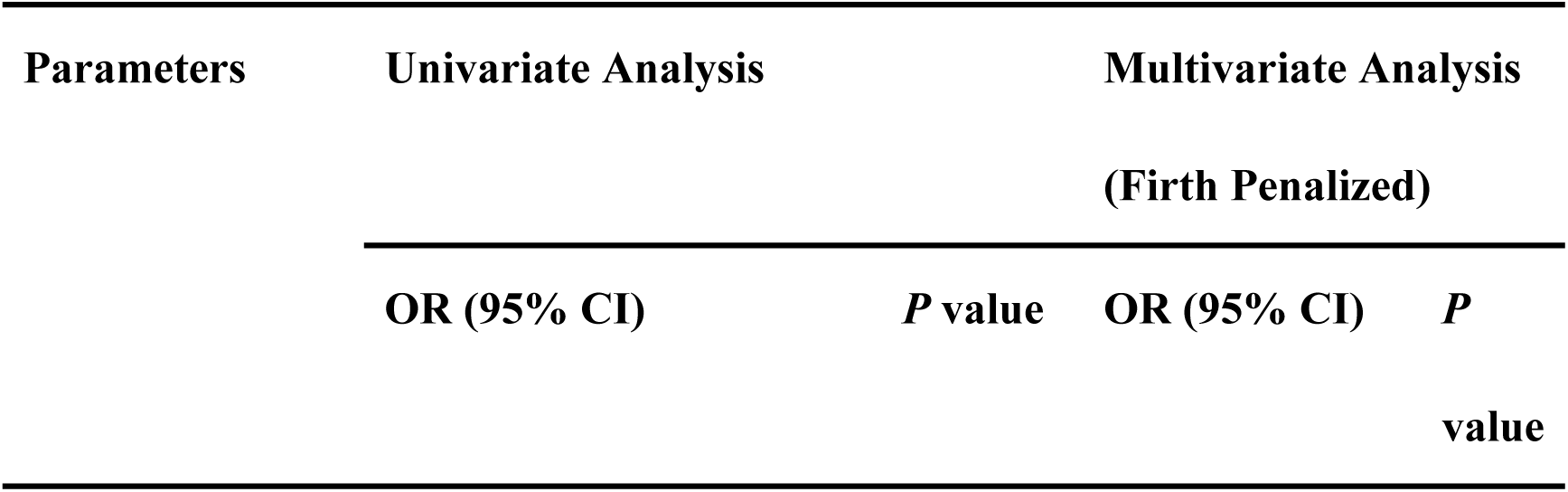
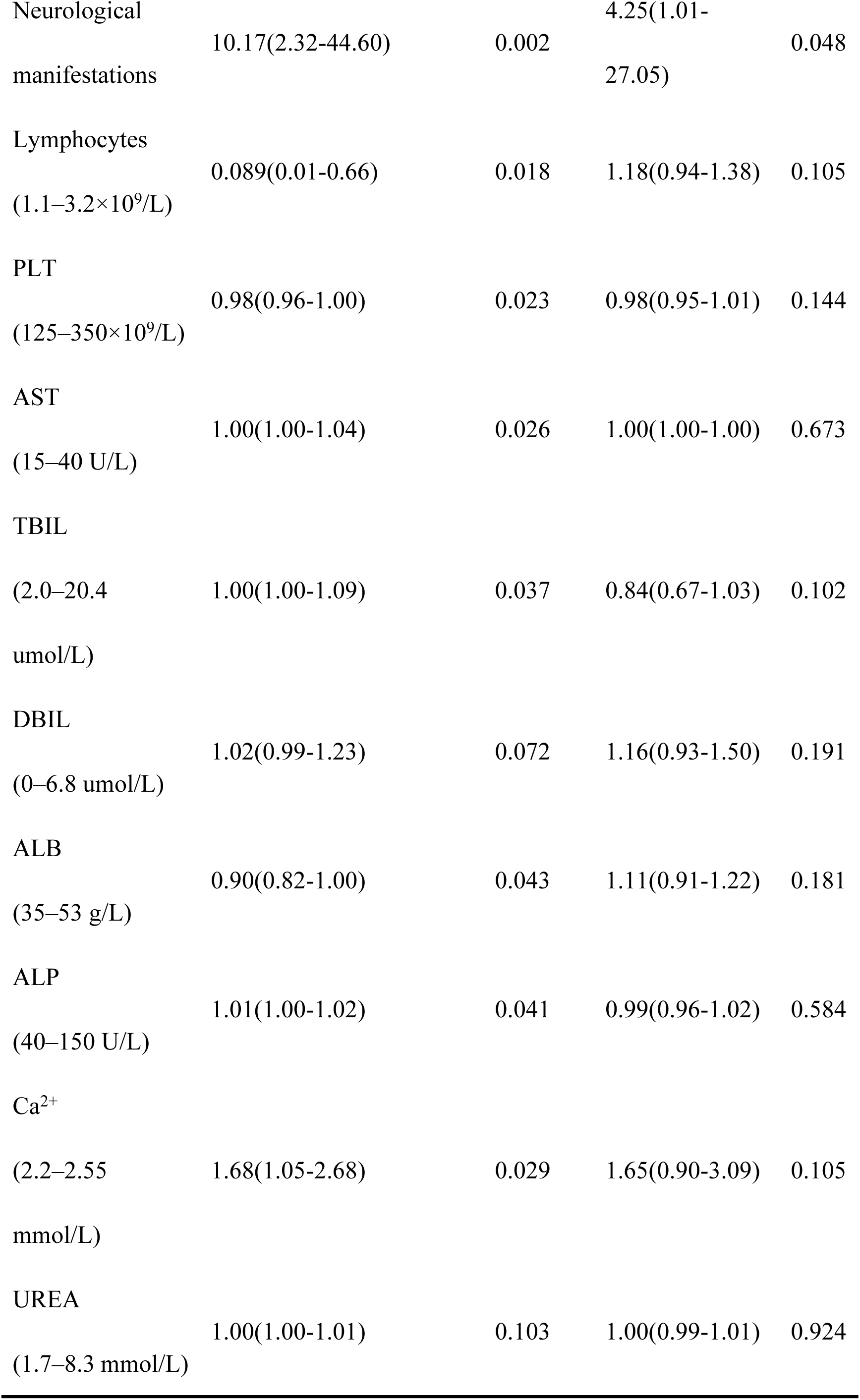
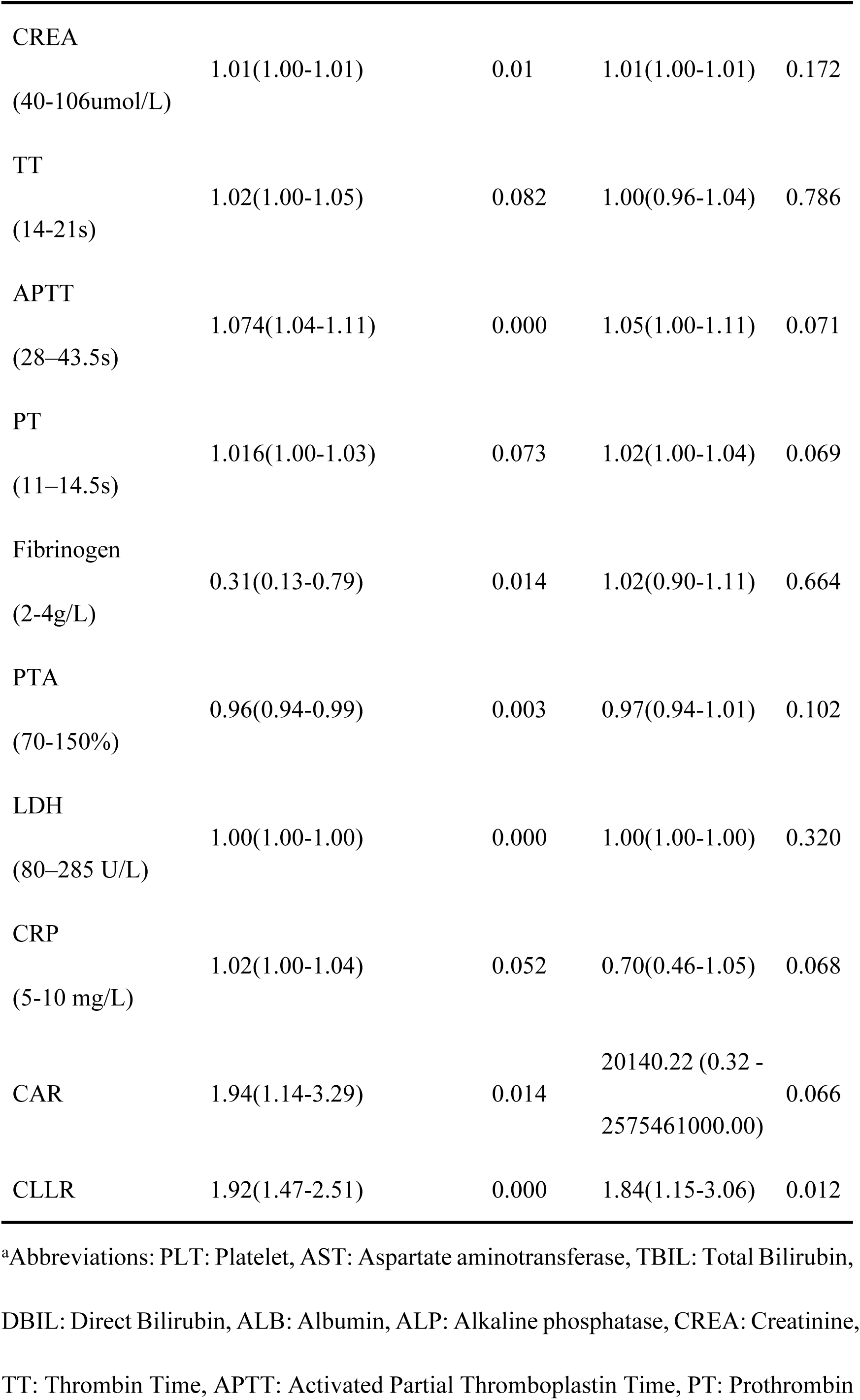
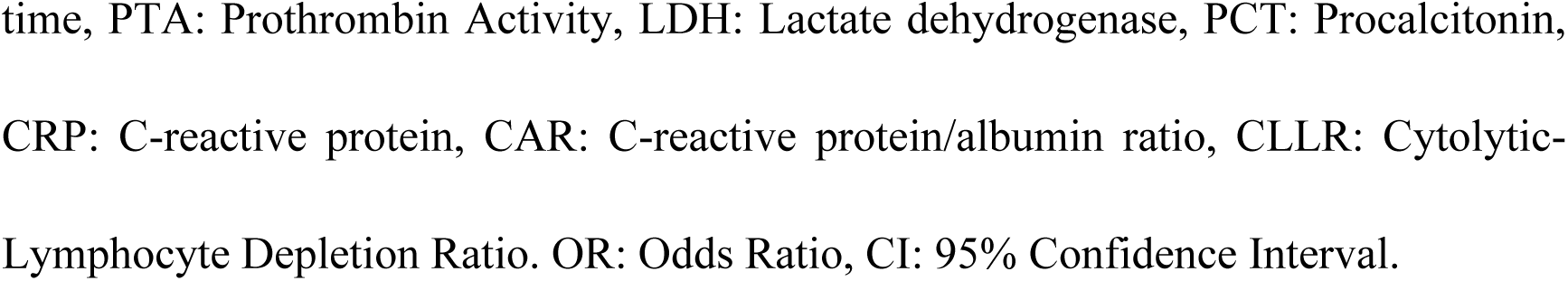
Risk factors associated with disease prognosis of patients with SFTS

### Diagnostic efficacy of sLLR for clinical outcomes in SFTS patients

ROC analysis was performed to compare predictive capacities between sLLR and CAR (Fig 1A). The AUC of the sLLR was 0.804, which was higher than that of CAR (DeLong’s test: ΔAUC=0.156, P=0.016). According to the maximum Youden index, the cut-off value of sLLR was calculated to be 1.457, with a sensitivity of 0.957 and a specificity of 0.595 (S1 Table).

**Fig 1.**
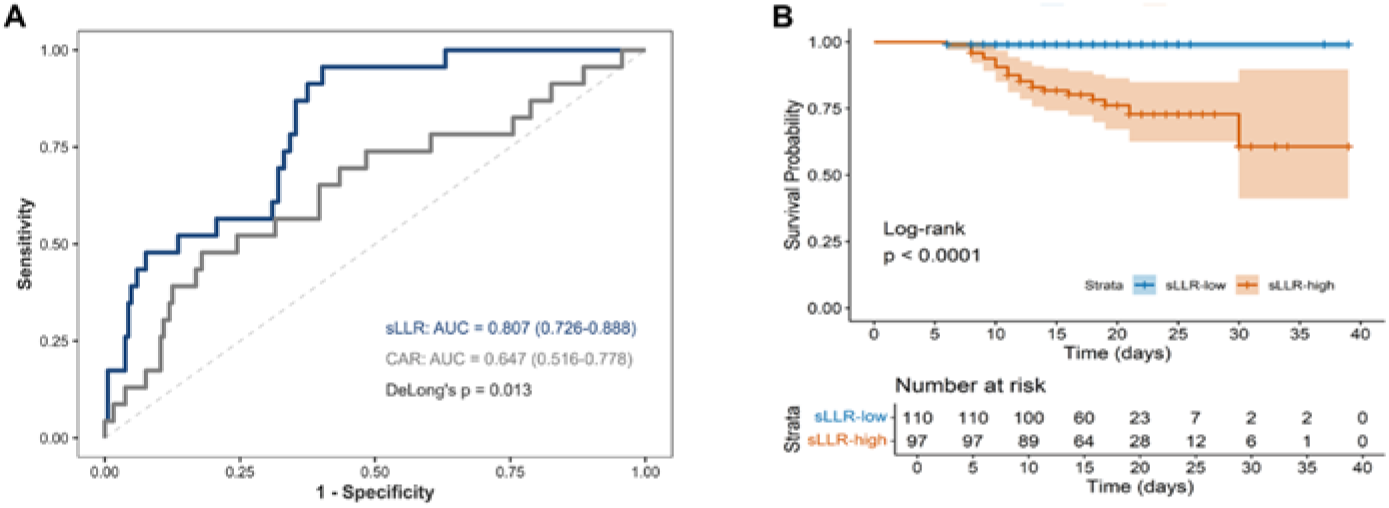
Predictive and prognostic value of sLLR in SFTS patients. A, Receiver operating characteristic (ROC) curves comparing the SLLR and CAR for mortality prediction in SFTS patients. B, Kaplan–Meier survival curves stratified by SLLR in SFTS patients.

We found that patients in the deceased group had higher sLLR than those in the survival group (S1A Fig). According to sLLR’s cutoff value (1.457), all patients were stratified into sLLR-low and sLLR-high subgroups. The mortality rate in the sLLR-high group was dramatically elevated compared to the sLLR-low group (S1B Fig).

According to Kaplan–Meier survival curve analysis, SFTS patients with higher sLLRs had lower cumulative survival than those with lower sLLRs (Log-rank, P < 0.001) (Fig 1B).

### Model Development and Validation

The predictive model, developed through Firth penalized regression, incorporated three independent mortality predictors. Multicollinearity was negligible (VIF <1.2 for all variables), and linearity assumptions were validated via Box-Tidwell analysis (P = 0.28 for sLLR and APTT transformations). The model demonstrated adequate goodness-of-fit (likelihood ratio test: deviance = 40.96, P < 0.001). This prediction model is visualized using a nomogram (Fig 2), which established a 220-point risk stratification scale with corresponding mortality probabilities ranging from 0% to 70%. The nomogram stratified patients into low-, intermediate-, and high-risk groups. Total scores stratified patients into three risk categories:

**Low-risk (<100 points):** 10% mortality.
**Intermediate-risk (100–152 points):** 50% mortality.
**High-risk (>152 points):** 70% mortality.

**Fig 2.**
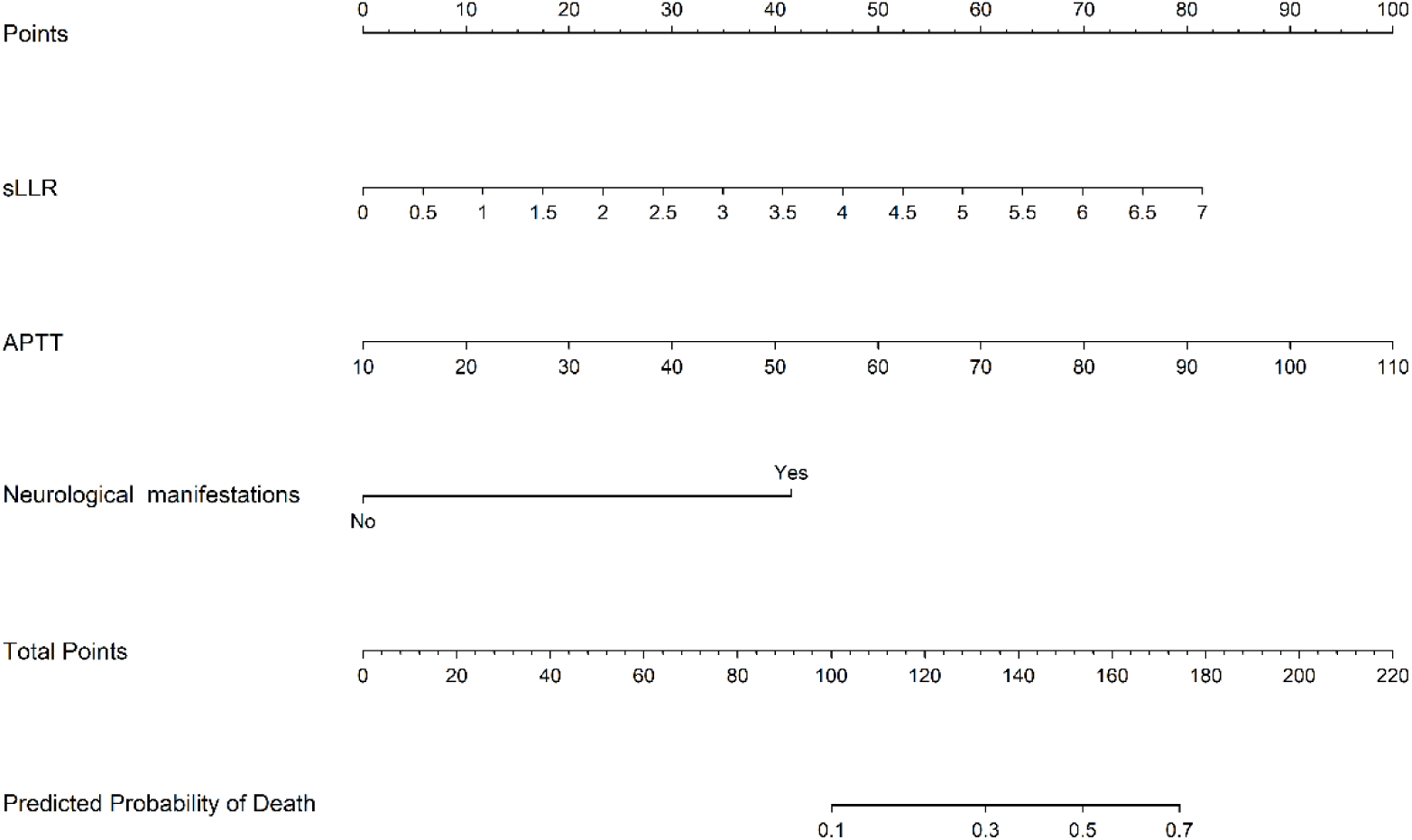
Nomogram for mortality risk stratification in SFTS patients based on the lactate dehydrogenase (LDH)/Lymphocyte ratio (SLLR), activated partial thromboplastin time (APTT), and neurological manifestations.

Each 20-point increase doubled mortality risk (aOR = 2.25, 95% CI 1.63–3.11; P < 0.001), providing risk stratification for clinical decision-making.

Internal validation through 1,000 bootstrap resamples showed robust predictive accuracy, with AUC of 0.876 (Fig 3A) Calibration analysis revealed alignment between predicted mortality risks and observed outcomes (Brier score = 0.074; Hosmer-Lemeshow P=0.778), with calibration curves tracking the ideal diagonal (Fig 3B). DCA quantified clinical utility across threshold probabilities (1%–66%). The model provided superior net benefit compared to default "treat all" or "treat none" strategies (Fig 3C).

**Fig 3.**
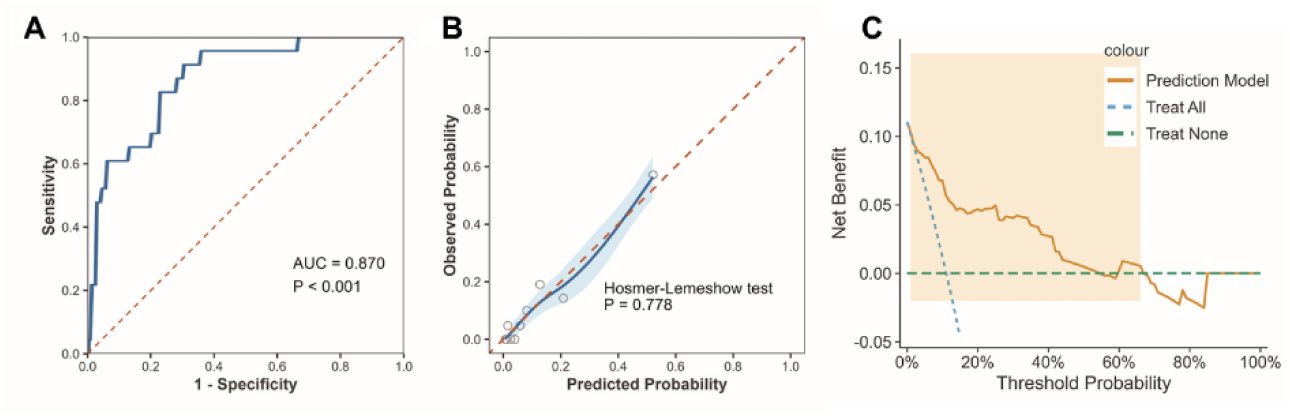
Internal validation and clinical utility of the sLLR-based prognostic model. A, Receiver operating characteristic (ROC) curve of the model evaluated through 1,000 bootstrap resamples, demonstrating robust predictive accuracy (AUC = 0.876, 95% CI: 0.777–0.923). B, Calibration curve comparing predicted mortality probabilities with observed outcomes. The model showed good alignment with the ideal diagonal (Brier score = 0.074; Hosmer-Lemeshow test, P = 0.778), indicating small deviation between predicted and actual mortality risks. C, Decision curve analysis (DCA) evaluating the clinical net benefit of the sLLR-based model across threshold probabilities. The model demonstrated superior clinical utility compared to "treat all" or "treat none" strategies within a threshold range of 1%–66%.

## Discussion

This study appears to be the first to explore the potential of sLLR as a predictor of mortality outcomes in patients with SFTS. The predictive effect of sLLR was evaluated and compared with the classic biomarker CAR. The results showed that sLLR could serve as an important independent predictor of death in SFTS patients, with a higher AUC and outperforming CAR. The nomogram based on sLLR also exhibited superior discrimination, calibration, and clinical utility.

We consider the predictive value of sLLR for the risk of death from SFTS from the perspective of the pathogenic mechanisms of the following pathogens and the pathological immune mechanisms of the host. The SFTSV evades innate immunity through its NSs protein, which inhibits the RIG-I signaling pathway by binding to the E3 ligase TRIM25, preventing RIG-I ubiquitination and blocking MAVS-TBK1-IRF3 activation[18]. The NSs protein also sequesters transcription factors like IRF3 and IRF7 in viral inclusion bodies, reducing type I interferon production and facilitating viral replication[18]. The viral Gn/Gc glycoproteins destabilize host cell membranes, leading to lactate dehydrogenase (LDH) release[19–21], which is significantly higher in deceased patients compared to survivors, correlating membrane damage with disease severity[12, 22]. Additionally, SFTSV activates the NLRP3 inflammasome, promoting lymphocyte depletion[23, 24]. The NSs protein interacts with NLRP3, driving inflammasome oligomerization and caspase-1 activation, which cleaves pro-IL-1β, causing CD4+ T cell pyroptosis and inhibiting bone marrow hematopoiesis[25, 26].

These mechanisms—elevated LDH and lymphocyte depletion—reflect a dynamic imbalance in immune response, contributing to disease progression and severity.

The CAR might be limited by the nonspecific inflammatory response of CRP and the fluctuations in nutritional and metabolic status associated with ALB. We found that sLLR could be an independent risk factor for assessing poor prognosis in SFTS patients through Firth penalized regression. The sLLR in the death group was 2.5 times higher than in the survival group, significantly greater than the 1.5-fold difference in CAR, suggesting that sLLR may have superior recognition ability for the specific pathological processes of SFTS[27, 28]. Additionally, we observed that sLLR exhibited a high sensitivity of 95.7%, significantly outperforming CAR’s 47.8%. This allows for early identification of high-risk patients requiring intensive care, enabling early intervention and dynamic assessment.

In our clinical model, APTT did not show statistical significance in multivariate analysis but was significantly associated with mortality risk in univariate analysis (P < 0.001), suggesting that APTT may have potential clinical significance in early mortality risk prediction. Previous studies have confirmed that APTT is closely related to the severity of disease in SFTS patients[29]. SFTS patients often have coagulation dysfunction, and prolonged APTT may be related to widespread inflammation and endothelial damage, which could increase the risk of mortality. The inclusion of APTT in our model reflects its impact on coagulation and inflammation in severe cases.

This predictive model provides an effective approach for risk stratification in SFTS patients. Its predictive stability, assessed through Bootstrap internal validation, demonstrates adequate performance for clinical applications. The key variable, sLLR, is derived from the ratio of LDH to LY, both of which are measured through routine tests, making the model cost-effective and easily implementable for rapid triage in primary care settings.

Our study has a few limitations. First, the single-center retrospective design may introduce selection bias. Although internal validation through Bootstrap was performed to reduce the risk of overfitting, multi-center prospective cohort studies are needed to validate the model’s generalizability. Second, the small number of death cases (n=23) may impact the stability of statistical inferences. Although Firth’s penalized regression corrected for small sample bias and optimized parameter estimates, this association requires validation in larger independent cohorts to confirm its reliability. Finally, the lack of dynamic monitoring of sLLR limits the understanding of its variation patterns. Future studies should integrate time-series analysis to clarify the temporal relationship between sLLR trajectories and clinical outcomes.

## Conclusion

This study identifies the standardized lactate dehydrogenase-to-lymphocyte ratio (sLLR) as a novel early warning indicator for fatal outcomes in hospitalized patients with SFTS. The nomogram incorporating sLLR, APTT, and neurological manifestations demonstrated excellent performance in predicting mortality outcomes in SFTS patients, offering a simple and intuitive triage tool for primary care physicians in resource-limited settings to make more informed clinical decisions.

## Data Availability

All relevant data are within the manuscript and its Supporting Information files.

## Acknowledgement

Zhihai Chen, Ling Lin, and Ruize Ma designed the study, had full access to all the data in the study, and took responsibility for the data’s integrity and analytical accuracy. Ruize Ma, Jingxia Wang and Ranran Wang contributed to collecting the clinical data. Ruize Ma, Hongxiao Wu, Ruihua Zhang and Yameng Mu organized the clinical data and conducted the statistical analysis. Ruize Ma wrote the article. All authors reviewed and approved the final version.

## Financial supports

This work was supported by the National Natural Science Foundation of China (No. 82072295).

## Author Contributions

Conceptualization: Ruize Ma, Zhihai Chen. Data Curation: Ruize Ma, Jingxia Wang Formal analysis: Xiao Yu Xue, Ranran Wang.

Methodology: Ruize Ma, Hongxiao Wu, Ruihua Zhang. Funding acquisition: Zhihai Chen

Project administration: Zhihai Chen, Ling Lin. Resources: Xiao Yu Xue, Ranran Wang, Yameng Mu. Supervision: Zhihai Chen.

Writing-original draft: Ruize Ma, Jingxia Wang.

Writing-review & editing: Ruize Ma, Jingxia Wang, Ranran Wang.

---

**S1 Fig.**
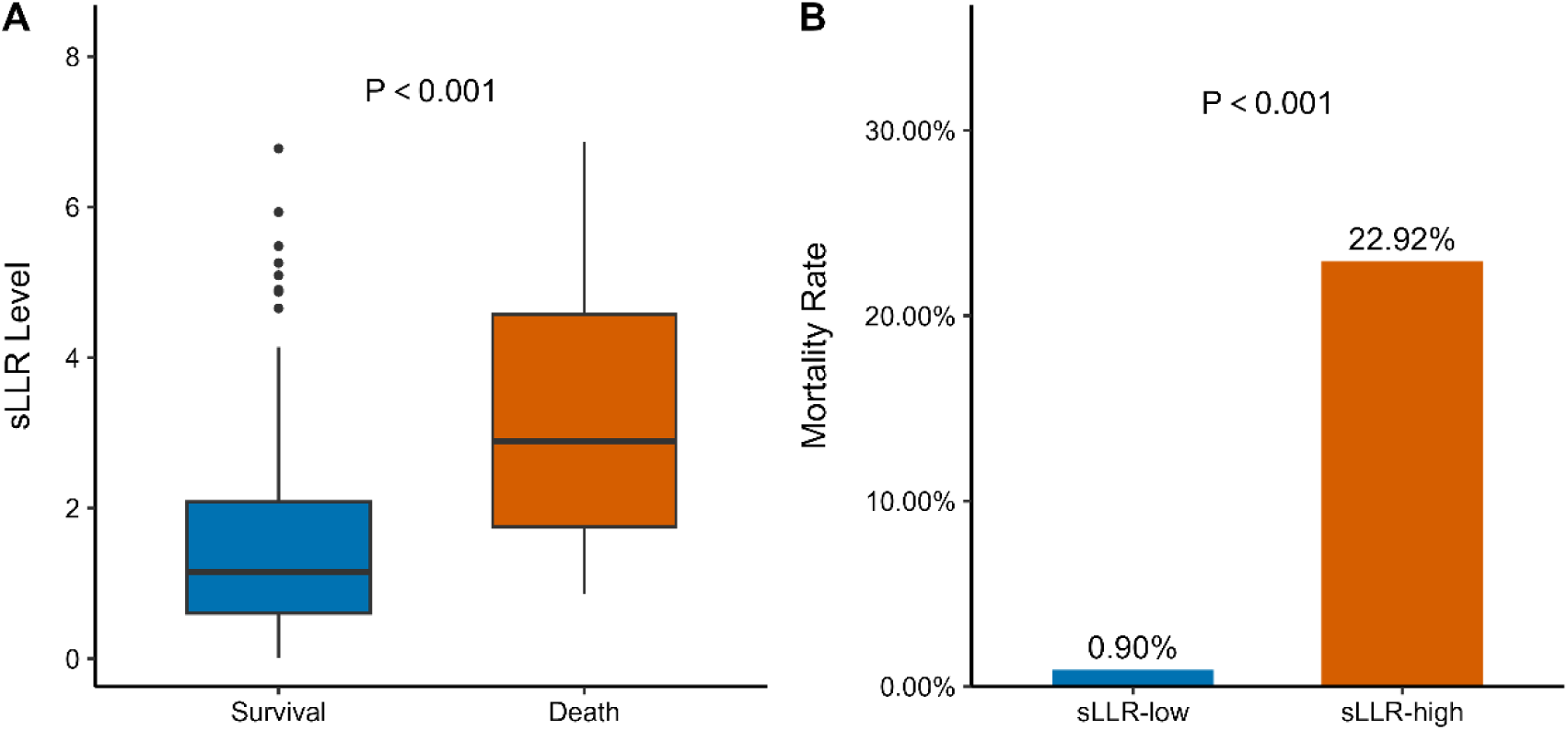
Association of sLLR with clinical outcomes in SFTS patients. A, Comparison of sLLR levels between survivors and non-survivors. Non-survivors exhibited significantly higher sLLR (median [IQR]: 2.88 [1.75–4.58]) compared to survivors (1.15 [0.60–2.09]; P < 0.001, Wilcoxon rank-sum test). B, Mortality risk stratification based on sLLR cutoff (1.457). The sLLR-high subgroup had a 31.88-fold increased odds of mortality compared to the sLLR-low subgroup (95% CI: 4.94– 1334.62; P < 0.001, Fisher’s exact test).

**S1 Table.** Predictive value of sLLR and CAR in predicting SFTS severity

| Parameters | AUC | Cutoff values | Sensitivity | Specificity | 95% CI | *p* value |
| --- | --- | --- | --- | --- | --- | --- |
| sLLR | 0.804 | 1.457 | 0.957 | 0.595 | 0.723-0.886 | < 0.001 |
| CAR | 0.648 | 0.362 | 0.478 | 0.822 | 0.517-0.779 | 0.021 |

*Note:* The cutoff points were selected by maximizing the sum of sensitivity and specificity.
Abbreviations: AUC, area under the ROC curve; CAR, C-reactive protein to albumin ratio; sLLR: lactate dehydrogenase/lymphocytes ratio. 95% CI, confidence interval.

